# Shaping sensory-association axis: genetics, transcriptomics, and relation to neuropsychiatry

**DOI:** 10.1101/2025.03.03.25323242

**Authors:** Bin Wan, Yuankai He, Varun Warrier, Alexandra John, Matthias Kirschner, Simon B. Eickhoff, Richard A.I. Bethlehem, Sofie L. Valk

## Abstract

The intrinsic functional organization of the human connectome can be captured along multiple spatial axes or gradients that differentiate sensory from association networks, visual from somatomotor networks, and control from default networks. These gradients demonstrate variability at the individual level, changing across the lifespan and varying across disorders. However, the biological basis of this variability is not fully understood. In this study, we integrated genetic, neuroimaging, and transcriptomic data using genome-wide association studies (GWAS) and twin-based heritability analysis of over 30,000 individuals to understand the contribution of genes to variability in functional organization gradients. Specifically, we identified five genetic loci involving 16 genes associated with the global organization of functional gradients that are enriched for lipid biosynthesis and energy metabolism. At the local regional level, we observe modest heritability scores across twin- and GWAS-based analyses of different samples, including adolescents, young adults, and middle-aged and older adults. Association areas are the most heritable. Importantly, the genes identified by GWAS do not overlap with those observed using post-mortem spatial transcriptomics. Lastly, while we did not observe genetic overlap between neuropsychiatric disorders and functional gradients, we found that regional gradient loadings are altered not only in neuropsychiatric disorders, but also in healthy individuals with polygenic risk scores for these disorders. Our findings highlight the complex interplay between genetic variation and intrinsic brain function. They offer new insights into the biological foundations of functional brain organization and its implications for neuropsychiatric vulnerability.

## Main

Functional gradients capture the spatial organization across the cerebral cortex, reflecting the inter-regional integration and segregation ^1–3^. They describe broad and major axes with functional relevance differentiating sensory/motor from more abstract cognitive functions ^1–3^. Moreover, these spatial axes of functional organization are spatially correlated with developmental timing ^4^, microstructure ^5,6^, and gene expression ^7,8^. Though these reports all point to a relationship between functional organization and its underlying neurobiology, most findings are at the group level and reflect spatial corrections, and little is known about the biological basis for individual differences in functional brain organization.

In particular, while these large-scale functional axes are consistently observed at the group level, substantial individual variability exists ^9,10^. For example, gradients vary over the lifespan ^11^, either in dominance of their axes, e.g., sensory-association and visual-somatomotor axes are shifted during childhood and adolescence ^12,13^ or integration and segregation of networks along gradient axes ^14^. Moreover, individual differences in gradients have been implicated in individual differences in creativity ^14^ and intelligence ^15^. Regional alterations along these axes are also observed in various psychiatric conditions such as autism ^17,18^, depression ^19^, and schizophrenia ^20,21^. However, the biological basis of this individual variability remains poorly understood.

Twin and family studies have demonstrated that functional brain networks exhibit low to moderate heritability, with estimates ranging from 20-40% for major resting-state networks ^22,23^, yet the genetic variants influencing intrinsic brain activity have remained largely undiscovered ^24^. Recent genome-wide association studies (GWAS) in large-scale datasets such as UK Biobank have begun to identify genomic loci associated with brain imaging phenotypes, including novel genetic loci associated with intrinsic functional signatures such as the default mode, central executive, and salience networks ^25^. Beyond regional measurements, recent work has explored the genetic basis of whole-brain network properties, identifying genes associated with global efficiency, local efficiency, and small-worldness of both functional and structural brain networks^26^. Emerging evidence indicates that functional gradients, capturing the hierarchical organization from unimodal sensory to transmodal association networks, are heritable and exhibit heterogeneous heritability patterns along the sensory-association axis ^27–29^, yet specific genetic loci associated with these gradient features remain to be identified through unbiased genome-wide approaches. Understanding the genetic architecture of functional brain gradients may provide critical insights into how genetic variation shapes the fundamental organizational principles of cortical function and their relationship to neuropsychiatric disorders.

Thus in the current work, we study the genetic basis of functional organization at the level of the individual using a multi-level approach.. We first constructed individual global features (spatial similarity to the canonical template) and regional loadings of the first three functional gradients differentiating sensory from association (G1), visual from sensorimotor (G2), and task-positive from task-negative (G3) networks, respectively. We then performed GWAS for them in the UK Biobank (middle-old adults). Second, we studied the regional heritability across age ranges to understand spatial variation in genetic influences with two additional twin-based data: Human Connectome Project (HCP, young adults), and Queensland Twin Adolescent Brain (QTAB, development). Third, transcriptomic mapping using the Allen Human Brain Atlas (AHBA) was analyzed to test whether identified genes by GWAS show expression patterns matching gradient organization. Finally, we used GWAS summaries and polygenic scores (PGSs) for 11 neuropsychiatric conditions, to understand how genetic predispositions for neuropsychiatry intersect the genetic basis of functional brain architecture.

## Results

### Phenotype construction

We generated the canonical gradient template using the HCP data, which forms previously reported sensory to association, visual-somatomotor, and modulation-representation axes ^3^ (**Fig. 1a**). As a template, HCP provides high-quality, multi-session, long-duration scans of resting-state functional magnetic resonance imaging (fMRI) in young adults. Then we aligned all individual gradients to the group-level HCP template. To do so, we computed the individual functional connectome based on atlas-wise resting-state functional connectivity and then reduced the dimensionality of its affinity matrix using diffusion map embedding. Procrustes alignment was used to rotate all individuals to the template, ensuring comparability. All the analyses were conducted based on a multimodal parcellation (MMP) that contained 180 pairs of bilateral symmetric brain cortical regions ^30^. The first three gradients accounted for over 50% (G1: 23.8%, G2: 19.1%, and G3: 15.8%, **Fig. 1a**) of the total variance in the functional connectome. Finally, we obtained comparable regional gradient loadings for each individual.

For the global feature, we calculated individual (dis)similarity scores (Pearson *r* and Euclidean distance) in the UKB with the high-quality HCP template to study individual variation in forming the whole sensory-association axis. The Pearson *r* reflects the linear relationship coefficient and Euclidean distance the absolute distance. Using these two alternative matrices can improve the ability of detecting significant SNPs. A higher similarity score indicates a typical sensory-association spatial pattern. We also found that lower similarity scores were correlated with higher age for G1 (*r* = -0.151, *P* < 0.001), G2 (*r* = -0.142, *P* < 0.001), and G3 (*r* = -0.107, *P* < 0.001).

### GWAS of the global feature-gradient similarities

We first aimed to investigate specific genes implicated in shaping individual variability of global functional brain organization patterns. To do so, we ran GWAS of the individual similarity scores before node-wise GWAS to uncover the related single-nucleotide polymorphisms (SNPs) using the GCTA ^31^ tool (https://yanglab.westlake.edu.cn/software/gcta/). We set minor allele frequency (MAF) to 1% to find the common variants, which include 9,116,883 remaining SNPs. We used the empirical significant threshold for *P* values (i.e., *P* < 5 × 10^-8^) and identified 3 loci containing 137 SNPs for G1 (Pearson *r*, **Fig. 1a**). Along the 3 loci (regional plot), the lead SNPs were rs1452628, rs3891783, and rs4880259. We observed 69 SNPs for G2 and 12 SNPs for G3 (**Supplementary Fig. S1**). Q-Q plots of the GWAS summary are shown in **Supplementary Fig. S2**. Detailed SNPs were reported in the **Source Data**. The heritability scores for the Pearson *r* were G1: 0.17 ± 0.019, G2: 0.10 ± 0.018, and G3: 0.12 ± 0.019. For Euclidean distance, we identified 5 loci (3 repetitive loci in Pearson *r*) containing 245 SNPs for G1 (**Fig. 2b**). Along the two non-repetitive loci (regional plot), the lead SNPs were 6:96893385:C:CAG and rs59375526. We observed 108 SNPs for G2 and 18 SNPs for G3 (**Supplementary Fig. S3**). The heritability scores for Euclidean distance were G1: 0.17 ± 0.019, G2: 0.11 ± 0.018, and G3: 0.09 ± 0.018.

We performed various robustness checks. In addition to Pearson *r* and Euclidean distance, we performed GWAS for cosine similarity as an alternative measure of similarity. We found similar Manhattan plots between Euclidean distance/Pearson *r* and cosine similarity (**Supplementary Fig. S4**). We also tested alternative gradient construction methods based on the top 50% of connections and the full functional connectome and performed GWAS for Pearson *r*. These analyses showed similar eigenmaps and consistent significant SNPs centered on chromosome 10 (**Supplementary Fig. S5 and S6**). Nonetheless, the top 10% has previously been shown to be more robust and offer better reproducibility and reliability ^3,9,10,32^.

We run a replication GWAS for the sensory-association axis (G1) using a new and independent European sample from the UKB (*N* = 14,471). As our Euclidean distance marker of similarity captured all significant loci in the main analysis, we performed comparisons between discovery and replication samples in the Euclidean distance feature of the sensory-association axis specifically. Of 245 genome-wide significant SNPs identified in the discovery sample, 244 (99.6%) were available in the replication sample. Of these 244 SNPs, 241 (98.8%) reached nominal significance (*P* < 0.05) in the replication sample, with near-perfect directional concordance (99.6%) and highly correlated effect sizes (Pearson *r* =0.98, *P* = 1.8e-171). The LDSC-based genetic correlation was positive (*r*_g_ = 0.27) but did not reach significance (*P* = 0.23), likely reflecting insufficient power in the replication sample (*h*^2^ = 0.085, SE = 0.055) rather than true genetic heterogeneity.

**Fig. 1:**
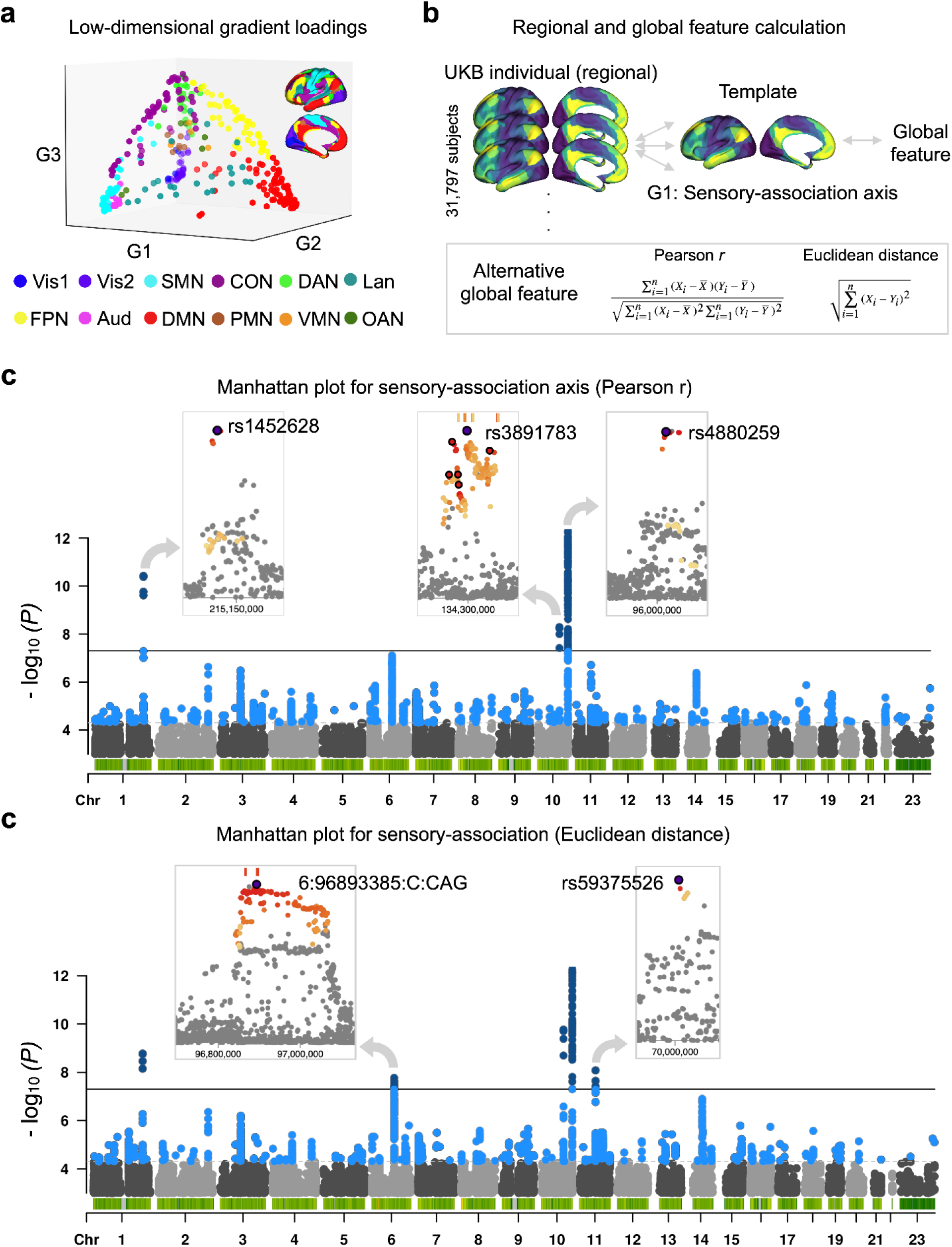
Genome-wide association studies (GWAS) on the similarity scores of Pearson *r* and Euclidean distance in G1. a. The low-dimensional space with colors coded by functional networks ^33^. Atlas-defined networks include primary visual (Vis1), secondary visual (Vis2), somatomotor (SMN), cingulo-opercular (CON), dorsal attention (DAN), language (LAN), frontoparietal (FPN), auditory network (AUD), default mode (DMN), posterior multimodal (PMN), ventral multimodal (VMN), orbito-affective (OAN). The three organization gradient axes were calculated from the function connectome ^27^ of the Human Connectome Project (HCP) ^34^. All UKB individuals were aligned to this HCP group-level template. We associated the UKB individual gradient maps with HCP group-level gradient maps to obtain (dis)similarity scores using Pearson *r* and Euclidean distance. Pearson *r* (**b**) uncovered three loci and Euclidean distance (**c**) uncovered two more loci in G1. Q-Q plots and GWAS summaries in G2 and G3 are shown in **Supplementary Figures** and **Source Data**.

### GWAS of the regional gradient loadings

To probe whether the GWAS observations of global gradient alignments at the individual level also reflect local variation in functional organization, we evaluated node-wise GWAS. The summaries with SNPs *P*-value < (5e-8)/360 for each gradient axis were given in **Source Data**. We then used principal component analysis (PCA) on the region-wise GWAS to obtain the multivariate GWAS output (**Fig. 2a**). It showed only one loci led by rs45411941 for G1. The genetic correlation between multivariate and similarity approaches was *r_g_* = 0.38 with *P* = 0.003. Afterwards, we plotted the region-wise significant SNPs for G1 in **Fig. 2b**. Briefly, we observed 4,654 significant SNPs for all regions along G1, of which 1,429 unique SNPs were identified for influencing different regions. For example, there were 44 SNPs from the 2nd chromosome, of which rs2863957 (maximum influencing SNP) influenced 31 brain regions including somatomotor areas. Likewise, we discovered 279 significant SNPs (183 unique) along G2 and 3,071 significant SNPs (733 unique) along G3. The most frequent SNPs were G1: rs2863957 influencing the somatomotor areas, G2: rs7632427 influencing Peri-Sylvian and visual areas, and G3: rs199993536 influencing somatomotor areas.

In addition, we found 4 SNPs (rs6540873, rs796226228, rs1452628, and rs10494988) in G1 global similarity also influencing regional loadings in the L_23d (the left middle cingulate cortex); 5 SNPs (rs11819412, rs4880259, rs4880396, rs200898494, and rs11146395) in G2 global similarity also influencing regional loadings in the R_FOP1 (the right frontal operculum); no SNPs in G3 global similarity.

**Fig. 2:**
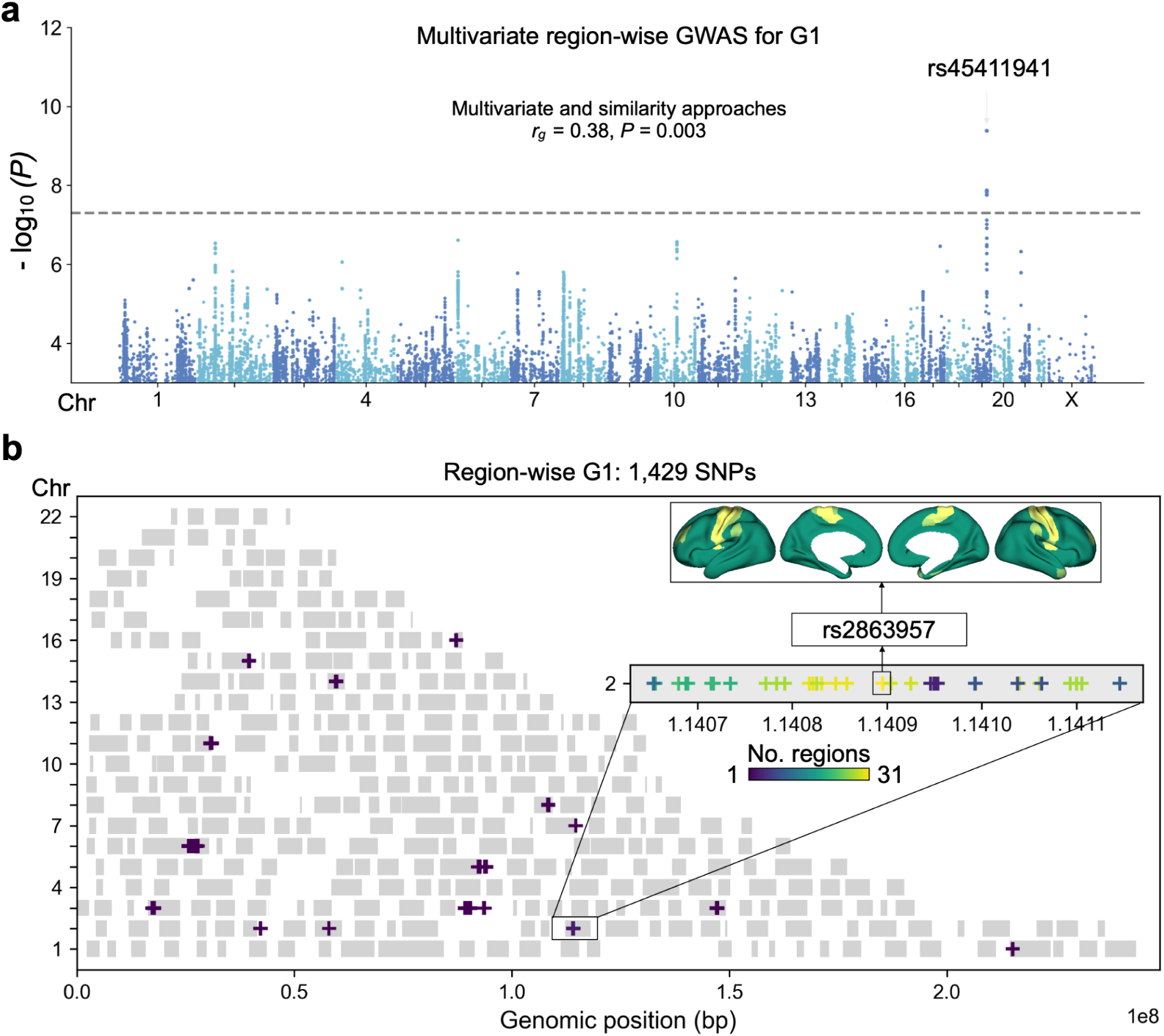
Node-wise GWAS on the gradient loadings. We performed 1,080 genome-wide association studies (GWAS) and discovered 4,654 significant single nucleotide polymorphisms (SNPs) for all regions along G1. Of these, 1,429 were unique and influenced different regions. Similarly, we identified 279 significant SNPs (183 unique) along G2 and 3,071 significant SNPs (733 unique) along G3. We then performed a multivariate principal component analysis (PCA) ^35^ on the region-wise results (**a**). We plotted “how many regions are the SNPs associated” along genomic position (**b**). For instance, 44 SNPs were found on the second chromosome, and rs2863957 influenced 31 brain regions, including somatomotor areas, along G1. The detailed regional and gradient annotations for each SNP can be found in the **source data**.

### Heritability of regional gradient loadings across three datasets

Following, we aimed to evaluate the heritability of regional loadings of functional gradients, to understand the portion of variance across individuals to be attributable to genetic factors. To do so, we analyzed data from twin-based and SNP-based cohorts. Utilizing the genotype information from UKB and pedigree information in HCP and QTAB, we calculated the heritability (*h*^2^) in each sample (detailed methods see **Methods**). Among G1, G2, and G3, dorsolateral prefrontal and posterior temporal regions showed a relatively moderate heritability (*h*^2^ > 0.2) and sensory areas showed a relatively low heritability (*h*^2^ < 0.01) in the UKB (**Fig. 3a**). It also showed similar results in twin-based HCP (**Fig. 3b**) and QTAB (**Fig. 3c**). Detailed heritability scores, standard errors, and *P*-values are shown in **Source Data**. For G1, the spatial correlations (Pearson) were *r* = 0.209 between UKB and HCP, *r* = 0.293 between UKB and QTAB, and *r* = 0.330 between HCP and QTAB, all *P*_variogram_ < 0.03. For G2, the correlations were *r* = 0.237 between UKB and HCP, *r* = 0.334 between UKB and QTAB, and *r* = 0.247 between HCP and QTAB with all *P*_variogram_ < 0.02. For G3, the correlations were *r* = 0.220 (*P*_variogram_ = 0.013) between UKB and HCP, *r* = 0.138 (*P*_variogram_ = 0.159) between UKB and QTAB, *r* = 0.185 (*P*_variogram_ = 0.016) between HCP and QTAB, **Fig. 3d**. These heritability estimates indicate that genetic factors explain modest amounts of individual variance in gradient organization, with the remaining variance likely attributable to environmental factors, measurement error, and gene-environment interactions. The moderate heritability combined with high spatial correlation across datasets suggests that while genetic influences on gradients are modest in magnitude, they are spatially consistent across the lifespan. Notably, higher heritability in association cortices compared to sensory regions may reflect either stronger genetic constraints on higher-order cognitive functions or greater environmental plasticity in sensory systems during development.

We also observed correlations of heritability across gradients (**Fig. 3d**). In the UKB, the spatial correlations (Pearson) were *r* = 0.453 between G1 and G2, *r* = 0.543 between G1 and G3, and *r* = 0.399 between G2 and G3, with all *P*_variogram_ < 0.001. In the HCP, spatial correlations (Pearson) were *r* = 0.359 between G1 and G2, *r* = 0.335 between G1 and G3, and *r* = 0.217 between G2 and G3, with all *P*_variogram_ < 0.02. In the QTAB, the correlations were *r* = 0.502 (*P*_variogram_ < 0.001) between G1 and G2, *r* = 0.302 (*P*_variogram_ < 0.001) between G1 and G3, and *r* = 0.126 (*P*_variogram_ = 0.197) between G2 and G3. Overall, our findings demonstrate reproducible spatially-correlated heritability patterns across datasets spanning different age groups, suggesting that genetic factors moderately influence gradient organization throughout the lifespan.

**Fig. 3:**
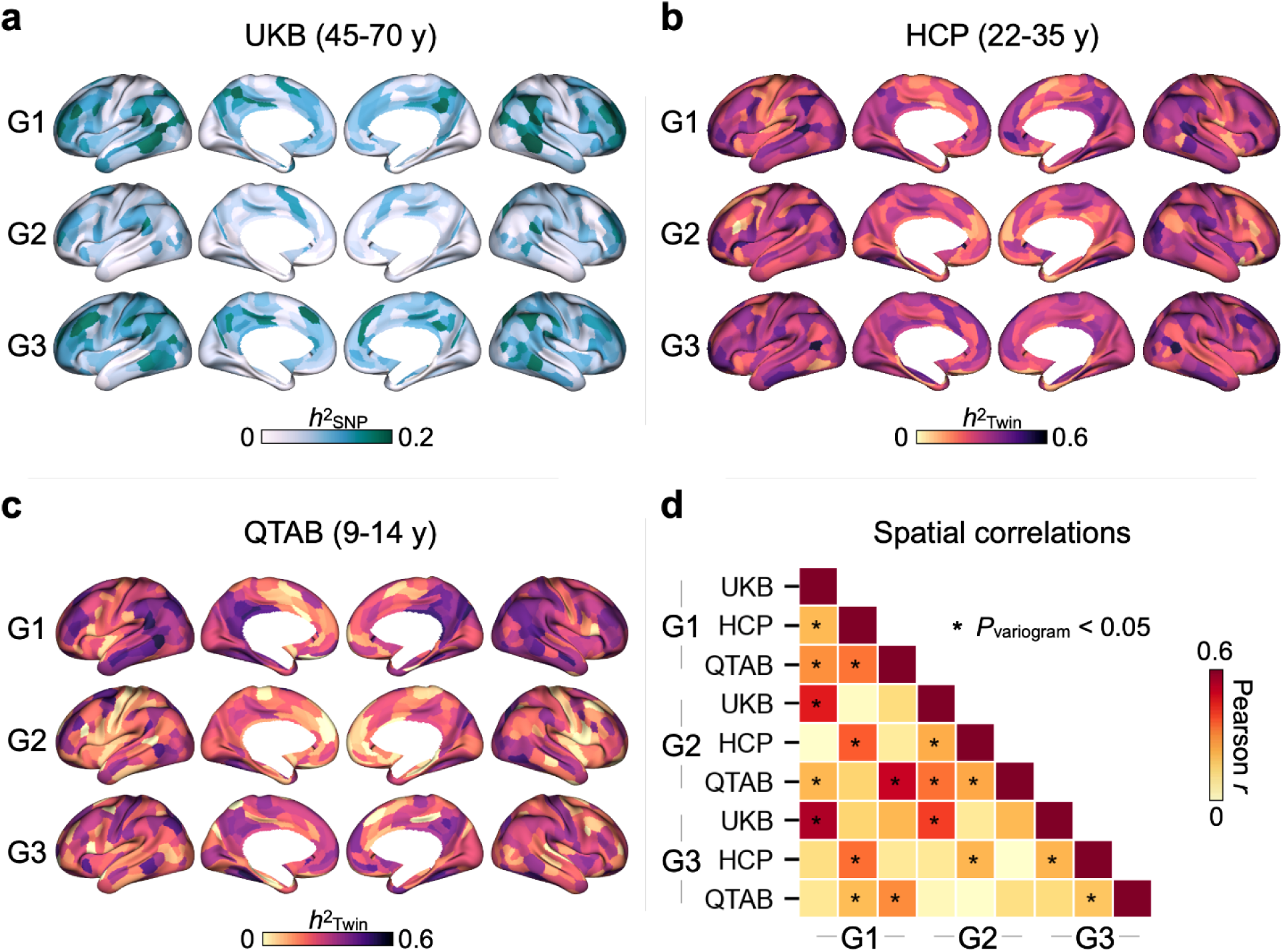
Heritability (*h*^2^) maps in different datasets. We used individual gradients and pedigree/genotype information to calculate the heritability (*h*^2^) per region per gradient for the single-nucleotide polymorphisms (SNPs)-based UKB (**a**), twin-based HCP (**b**), and twin-based QTAB (**c**). **d**. Spatial correlation between each two heritability maps. Spatial autocorrelation is considered to permute the maps using the geodesic distance variogram and *P*_variogram_ values are obtained based on 1000 spatial autocorrelation surrogate maps.

### Overlap between genetic loci and gene expression related to the global feature

Next, we wanted to understand how the genetic loci identified for global functional organization markers relate to cortical gene expression. This approach would help uncover the molecular pathways through which genetic variation influences functional gradient organization. First, we used the functional mapping and annotation (FUMA) ^36^ tool (https://fuma.ctglab.nl/) to map the nearest genes based on the GWAS results (maximum distance: 10kb) as well as MAGMA gene-based analysis ^37^. We mapped 6 genes in total for the Pearson *r* using positional mapping (*FAM200A*, *NPC3L*, and *PLCE1*) and MAGMA (*GNA12, FHL5,* and *AMZ1*), shown in **Supplementary Fig. S8**. Regarding Euclidean distance, in addition to the overlapping genes, we mapped 10 genes including positional mapping (*FUT9, TBC1D12*, *HELLS*, *CYP2C9*, *CYP2C8*, *SLC35G1*, and *RP11-805J14.3)* and MAGMA (*UFL1*, *FHL5*, and *ANO1*), shown in **Supplementary Fig. S9**. We also performed summary data-based Mendelian randomization (SMR) using four quantitative trait locus (QTL) datasets from human brain tissues: adult prefrontal cis-acting chromatin accessibility QTLs (PFC_caQTL), PsychENCODE expression QTL (PsychENCODE_eQTL), PsychENCODE isoform-level QTL (PsychENCODE_isoQTL), and PsychENCODE splicing QTL (PsychENCODE_sQTL). While no significant genes were identified in the three PsychENCODE datasets, we detected one significant gene, *NDAFU12*, in PFC_caQTL (see **Supplementary Fig. S10 and S11**). Note that our main metric of gradient similarity is a global feature and not a local prefrontal feature. Thus, it is not possible to verify spatial associations across the entire cortex when using SMR results instead of positional mapping and MAGMA.

To understand how the transcriptomics of these genes show a spatial relationship with functional gradients, we extracted gene expression data from the AHBA (**Fig. 4a**). AHBA contains approximately 500 brain locations per hemisphere across six donors with two donors having data from both hemispheres and four donors only having left hemisphere data ^38^. When mapping the brain locations of probes onto the MMP atlas, there were 170 (of 180) regions available in the left hemisphere and 127 (of 180) regions available in the right hemisphere. The gene expression maps are offered by the ABAGEN ^38–40^ and were cleaned using the ENIGMA toolbox ^41^. We imputed the missing values using the variogram algorithm based on the central coordinates of the parcels and used imputed data for any downstream analyses. Among the 16 genes, we discovered 10 available in AHBA. The 10 gene expression maps are displayed in **Fig. 4b**.

Our analyses indicate that functional organization reflects a multivariate genetic architecture rather than single-gene effects. To quantify how the joint expression pattern of these 10 genes shapes cortical gradient topography (G1: sensory-association, G2: visual-somatomotor, G3: modulation-representation), we modeled each gradient’s spatial profile (360 regional loadings) as a linear combination of the regional expression levels of all 10 genes using ordinary least squares regression. We used (n) significant genes (*P* < 0.05) in OLS to compare the randomized resampling (15,000 samples) combination using the same number (n) of genes shown as null distribution. We defined its specificity as the percentage of how many of the samples exceeded the ‘practical’ *R*^2^ (OLS with significant genes). *R*^2^ was 0.219, and *R*^2^ was 0.206 with six significant genes and specificity = 0.904 for G1 (**Fig. 4c**). Regarding G2, *R*^2^ was 0.189 and *R*^2^ was 0.177 with four significant genes with specificity = 0.978 (**Fig. 4d**). Regarding G3, *R*^2^ was 0.083 and *R*^2^ was 0.067 with three significant genes with specificity = 0.973 (**Fig. 4e**). This indicates that the GWAS-informed gene set reaches an excellent fit at the transcriptome level.

Finally, as the above analyses were informed by GWAS hits, we also tested a purely data-driven approach to understand how a combination of transcriptomic maps could describe three gradients simultaneously based on spatial associations (Y: 360 gradient loadings, X: 360 expression values by #significant genes). We used a total of 15,634 genes as the input, then used elastic net (lasso ratio: 0.1-1) as the estimator to sparsify features, and finally detected which of them could be selected to predict three gradients (**Fig. 4f**). We detected 46 to 5 gene expression maps using a lasso (L1) ratio from 0.1 to 1 (**Fig. 4g**). The weight of the selected genes was displayed as a word cloud. The top 6 genes were: *COL27A1*, *COL8A2*, *ADAD2*, *AMDHD1*, *CCDC158*, and *CD6*. The full list of the genes is provided in **Source Data**. Most of the parameters suggested an excellent specificity for G1, G2, and G3. We then used the same number as GWAS-informed genes for each gradient (i.e., lasso ratio = 0.5, 10 genes in total, **Fig. 4h**). It reached a maximum (> 0.98) specificity (8 genes for G1, 6 genes for G2, and 4 genes for G3) but the genes selected were totally different from GWAS.

Importantly, despite both achieving high specificity in modeling gradient spatial patterns, we observed no overlapped genes between GWAS and data-driven AHBA approach. This discordance likely reflects fundamental methodological differences: GWAS identifies variants associated with individual differences in gradient organization across ∼30,000 individuals, while AHBA analysis examines spatial correlation between averaged gene expression and group-level gradients across ∼300 cortical locations from 6 post-mortem brains. The GWAS approach captures genetic variation that influences gradients through developmental, physiological, or plastic mechanisms, whereas AHBA identifies genes whose spatial expression patterns simply correlate with gradient topography, without establishing causality or relevance to individual differences. This suggests these approaches capture complementary but distinct aspects of the gene-gradient relationship, with GWAS findings more directly relevant to understanding heritable individual variation.

**Fig. 4:**
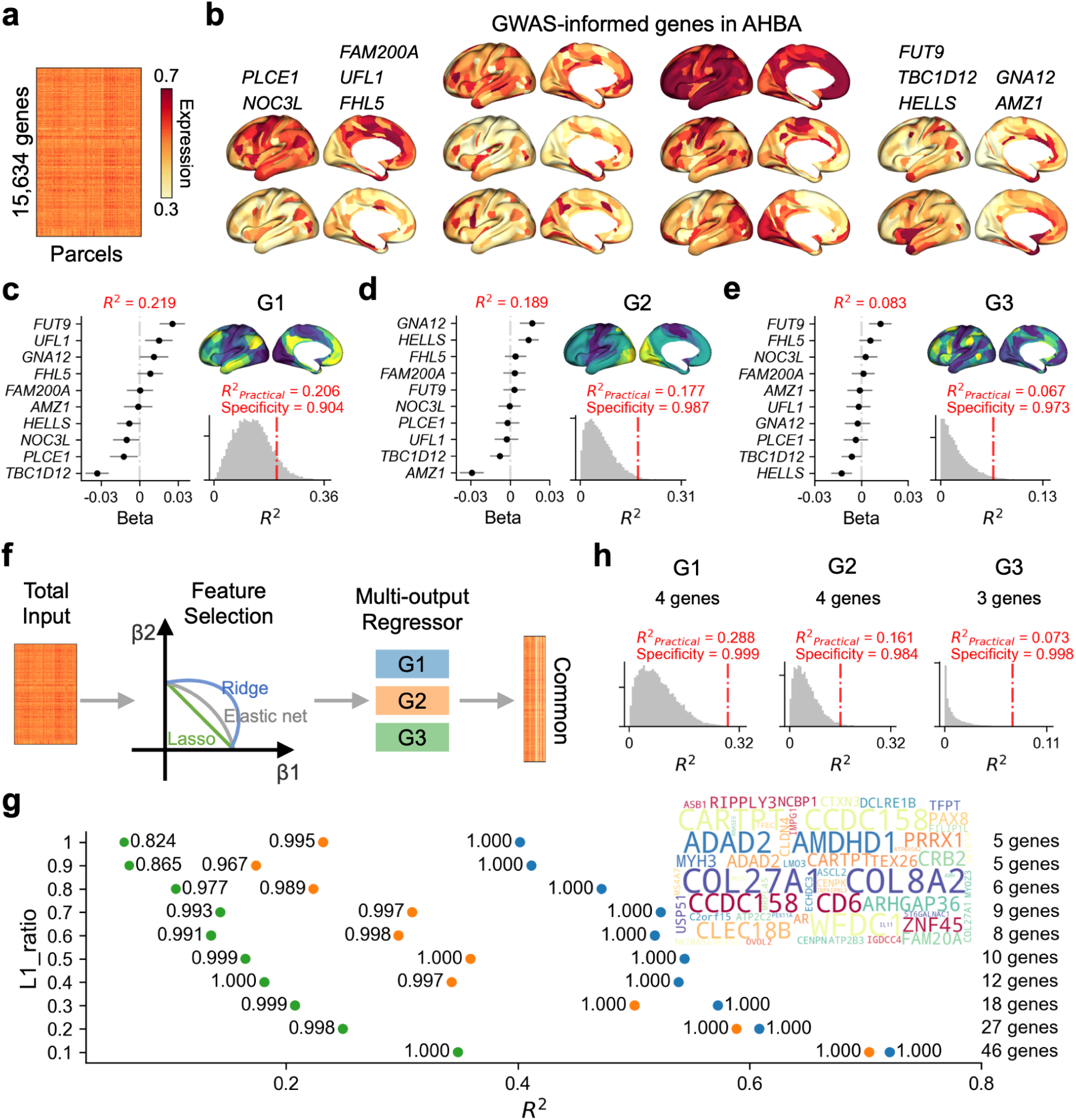
Genetic validation of gradients using transcriptomic data. **a** Shows gene expression extracted from the Allen Human Brain Atlas (AHBA) with 15,634 genes by 360 parcels (297 available). **b** Shows genes mapped from the GWAS summary using the FUMA tool, including positional mapping (maximum distance: 10kb) and MAGMA results. **c** The beta values of the six genes with a 95% confidence interval (CI) using an ordinary least squares (OLS) model to fit the G1 map as well as how many resampling samples (15,000) below the practical model (four significant genes, *P* < 0.05) for G1. **d** OLS and resampling (four significant genes, *P* < 0.05) for G2. **e**. OLS and resampling (three significant genes, *P* < 0.05) for G3. **f** The data-driven approach using an elastic net to select seven common contributing genes for the three gradients. **g** Tests different lasso (L1) ratios for detecting the common genes. The number next to each dot indicates the specificity of the selected gene set. The word cloud indicates the frequency of the selected genes (separate gradients in **Supplementary Fig. S12**). **h** Shows bootstrapping for three gradients after the data-driven selection (L1_ratio = 0.5 for G1, G2, and G3).

### Genetic correlation between functional gradients and macro- and micro-structural brain phenotypes

Next we aimed to understand whether the observed genes linked to individual variability of global functional organization related to brain structure. To explore the wider spectrum of brain structural and disease correlates of functional gradients, we used linkage disequilibrium (LD) scores ^42,43^ to estimate the SNP-based genetic correlations between functional gradients and genetic profiles in 13 macro- and micro-structural brain phenotypes based on previous GWAS summaries ^44^, further detailed in the **Methods**.

We identified stable genetic correlations for all three primary functional gradients and brain microstructural phenotypes derived from diffusion-weighted imaging (DWI) reflecting microstructural integrity ^44^ based on the Pearson *r* similarity metric (**Fig. 5a**). In particular, isotropic volume fraction (ISOVF, G1: *r_g_* = -0.373, *P*_FDR_ < 0.001; G2: *r_g_* = -0.307, *P*_FDR_ = 0.008; G3: *r_g_* = -0.366, *P*_FDR_ = 0.003), mean diffusivity (G1: *r_g_* = -0.294, *P*_FDR_ = 0.010; G2: *r_g_* = -0.254, *P*_FDR_ = 0.049) and orientation dispersion index (G1: *r_g_* = -0376, *P*_FDR_ = 0.004; G3: *r_g_* = -0.508, *P*_FDR_ < 0.001) were all negatively correlated with the functional gradients. We found similar genetic correlations for the Euclidean distance functional organization metric (**Fig. 5a**). Gradients were observed to be genetically inter-correlated amongst themselves (G1 and G2: *r_g_* = 0.873; G2 and G3: *r_g_* = 0.610; G3 and G1: *r_g_* = 0.735, **Fig. 5b**). In addition, the genetic correlations were also intercorrelated across Euclidean distance (dissimilarity) and Pearson *r* (similarity) with *r_g_* < -0.885 for G1 (**Fig. 5b**).

**Fig. 5:**
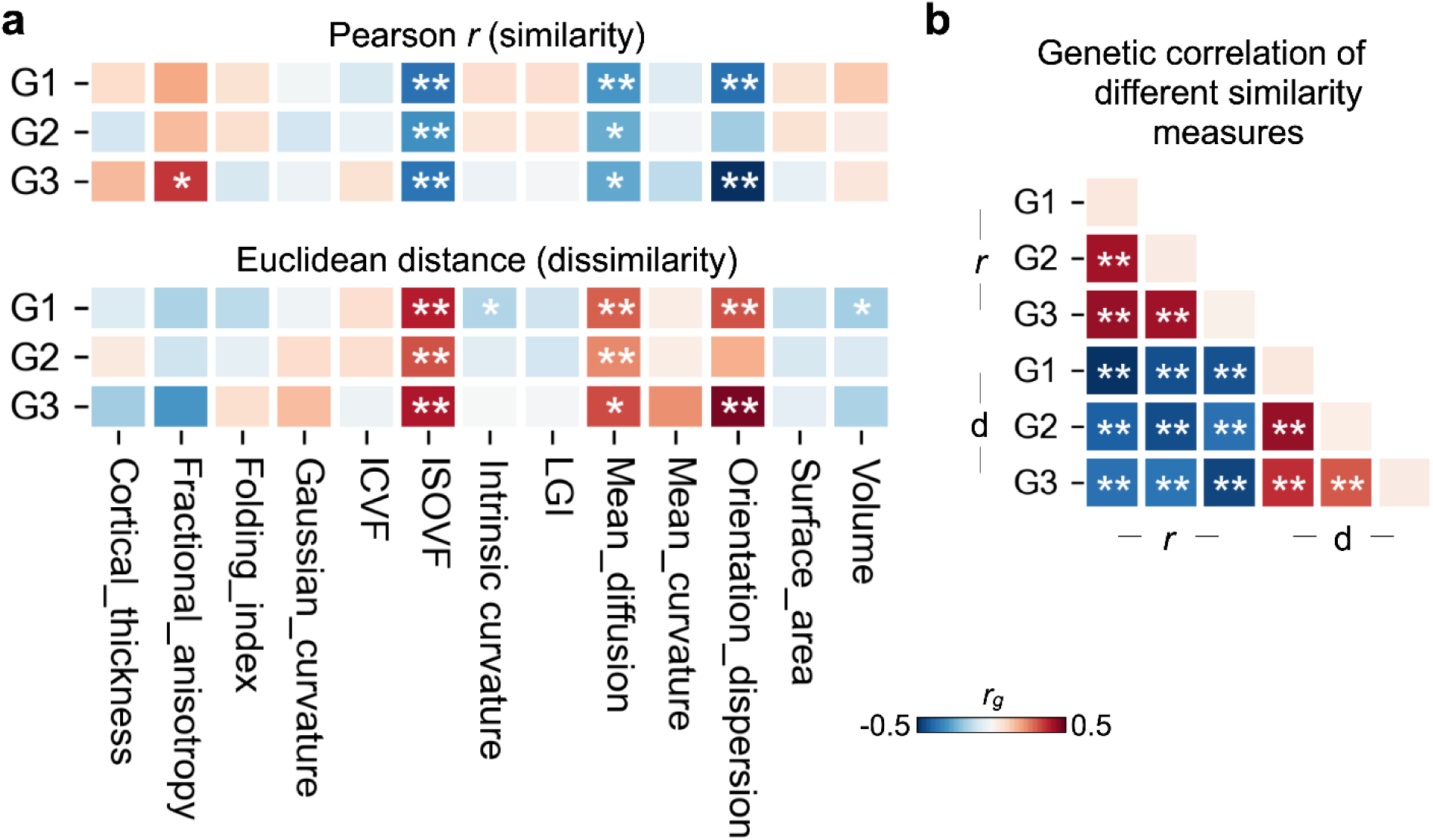
SNPs-based genetic correlations (*r*_g_) between functional gradients and 13 structural brain measures. **a** Shows genetic correlations for Pearson *r* (similarity) and Euclidean distance (dissimilarity) features. **b** Shows genetic correlations within gradients and between similarity measures. ICVF: intra-cellular volume fraction; ISOVF: isotropic volume fraction; LGI: local gyrification index; * and ** indicate *P* < 0.05 and *P*_FDR_ < 0.05.

### Functional annotations of the observed genes related to functional gradients

To understand how the observed genes related to global organization of functional gradients are expressed in the brain across development, we used BrainSpan (https://www.brainspan.org/) to map the averaged gene expression level from 8 weeks post conception (PCW) to 40 years after birth (in total 29 time points). This data has been integrated into FUMA. We evaluated gradient-related gene expression using the model in **Fig. 4b**. For example, the *t*-values returned for G1 by the OLS model were multiplied by the normalized expression score. The scores were then summed up and we looped it for the 29 development stages (**Fig. 6a**). We observed that G1-related genes were increasingly expressed after 13 years of age. G2- and G3-related genes were mostly expressed before birth. G2-related gene expression kept declining after birth whilst G3 remained stable in later life. The data points were smoothed by a Gaussian filter. We also performed this analysis for the AHBA-driven genes reported in the previous section. It showed that G1 expression remains stable but G2 increases expression of G2-related genes, and G3 expression initially decreases, but increases around pre-puberty (∼age 9) (**Fig. 6a**). Specific genes (in grey tones) did not show much variance in expression over time. Again there was little overlap between GWAS-based and spatial transcriptomics-based genes.

We then performed an enrichment analysis to understand the biological annotations of related genes using ShinyGO 0.81 ^45^. **Fig. 6b** shows 11 significant enriched terms for GWAS-informed genes. We used fold enrichment, a measure of the degree to which a particular term is overrepresented, in the genes of interest. These biological processings for GWAS-informed genes were centered on metabolic pathways and extended to lipid-related biosynthesis, acid-related metabolism, and serotonergic synapses (**Fig. 6c**). When performing enrichment analysis for the 10 AHBA-driven genes (L1_ratio = 0.5), we found there were two significant terms: histidine metabolism and protein digestion and absorption. There were no significant enriched terms using L1_ratio = 0.1, 0.2, 0.8, 0.9, and 1.

**Fig. 6:**
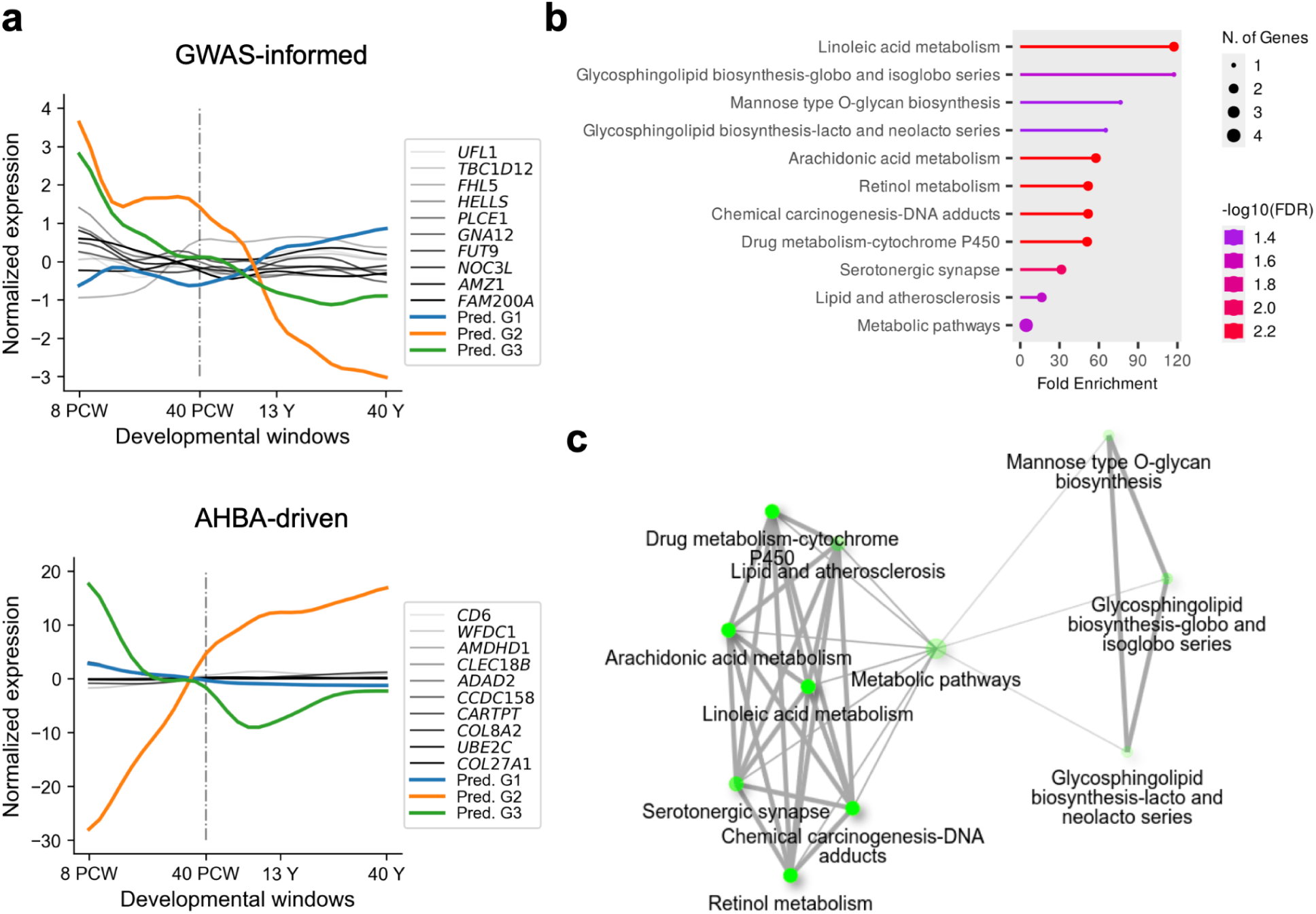
Developmental and functional annotations for the mapped genes. **a** Shows the cross-sectional trajectories of the GWAS-informed and AHBA-driven gene expressions, post-conception weeks (PCW), and years after birth (Y) data from *post-mortem* brain. Gaussian normalized scores with averaging across the brain tissues were used here. **b** Shows the enriched terms derived using ShinyGO version 0.81 ^45^ for the GWAS-informed genes. Fold enrichment is a measure of the degree to which a particular term is overrepresented in the genes of interest compared to a background set of genes. **c** Enriched significant terms visualized as a network with an edge threshold setting of 0.3 for the GWAS-informed genes.

### Association between regional gradients and neuropsychiatric PGSs

In the final step, we assessed the relationship between functional gradients and neuropsychiatric conditions using two complementary approaches. First, we tested whether the genetic signature of gradient organization (from our GWAS) directly overlaps with genes implicated in psychiatric disorders using genetic correlation analysis. This tests whether the same genetic variants influence both gradients and psychiatric risk. Second, we examined whether cumulative polygenic risk scores (PGS) for 11 disorders correlate with regional gradient loadings across individuals. This tests whether aggregate genetic liability relates to gradient patterns, even if individual variants differ. Using GWAS summaries of gradient similarity and each neuropsychiatric condition, we did not find any significant genetic correlation that survived multiple-testing correction (**Fig. 7a**), suggesting that the genetic signature found for global functional organization does not overlap with genes implicated in these conditions.

Alterations of regional gradient loadings have been observed in multiple psychiatric diseases ^17–20^, yet this genetic correlation misses some signals in regional loadings as we showed only a few overlapped SNPs for global and regional features in **Fig. 3**. To further test this, we constructed multivariate association using polygenic scores (PGSs) for 11 disorders in relation to regional gradient loadings. Sex, age, forty genetic principal components, head motion, and data site were regressed out for individual regional gradient loadings. Given the expected intercorrelation among the PGSs (**Fig. 7b**), suggesting transdiagnostic signatures ^46–48^, we conducted a partial least squares (PLS) analysis, with a machine learning pipeline, to uncover the overall multivariate association between regional gradients (360 x 30,716) and PGSs (11 x 30,716 x 3 gradients) in the UKB. We split the subjects into training and test samples 100 times and performed PLS in the training sample and transformed the test sample to study the brain-PGS first dimension correlation (**Fig. 7c**). We selected the model based on a balance between overfitting and underfitting (**Fig. 7d**). The first latent dimension suggested a positive correlation between PGS and gradient scores in the training sample (G1: *r* = 0.067, *P* = 5.64e-17; G2: *r* = 0.061, *P* = 3.48e-14; G1: *r* = 0.076, *P* = 2.83e-21; **Fig. 7d**). When studying the brain loadings (**Fig. 7e**), we observed a pattern differentiating central regions from frontal/occipital regions for G1, a pattern highlighting occipital regions for G2, and a pattern highlighting central sulcus for G3. When summarizing into 12 functional networks ^33^, we found the high averaged absolute loadings in auditory and language networks but low loadings in somatomotor, ventral multimodal, and primary visual networks. The regional correlation maps between gradients and PGS for each neuropsychiatric condition separately are shown in **Supplementary Fig. S13-15**. Here only few parcels survived the FDR correction (**Source Data**). Then, we correlated the brain loadings with the regional correlation maps to uncover the brain latent dimension was driven by which disorder (**Fig. 7f**). The SCZ PGS showed a strong correlation with the latent brain pattern (*r* = 0.829, 0.871, and 0.797 for G1-3), followed by BP (*r* = 0.669, 0.826, and 0.512 for G1-3) and PTSD (*r* = 0.500, 0.592, and 0.758 for G1-3). ADHD showed reverse correlation patterns for G2 (*r* = -0.293). The contrast between null genetic correlations (**Fig. 7a**) and significant PGS associations (**Fig. 7c-f**) indicates that psychiatric conditions and functional gradients do not share specific genetic loci, but rather that cumulative genetic risk across many variants relates to gradient organization. This suggests a complex polygenic architecture where numerous small-effect variants jointly influence both brain organization and psychiatric vulnerability through partially overlapping but distinct biological pathways.

**Fig. 7:**
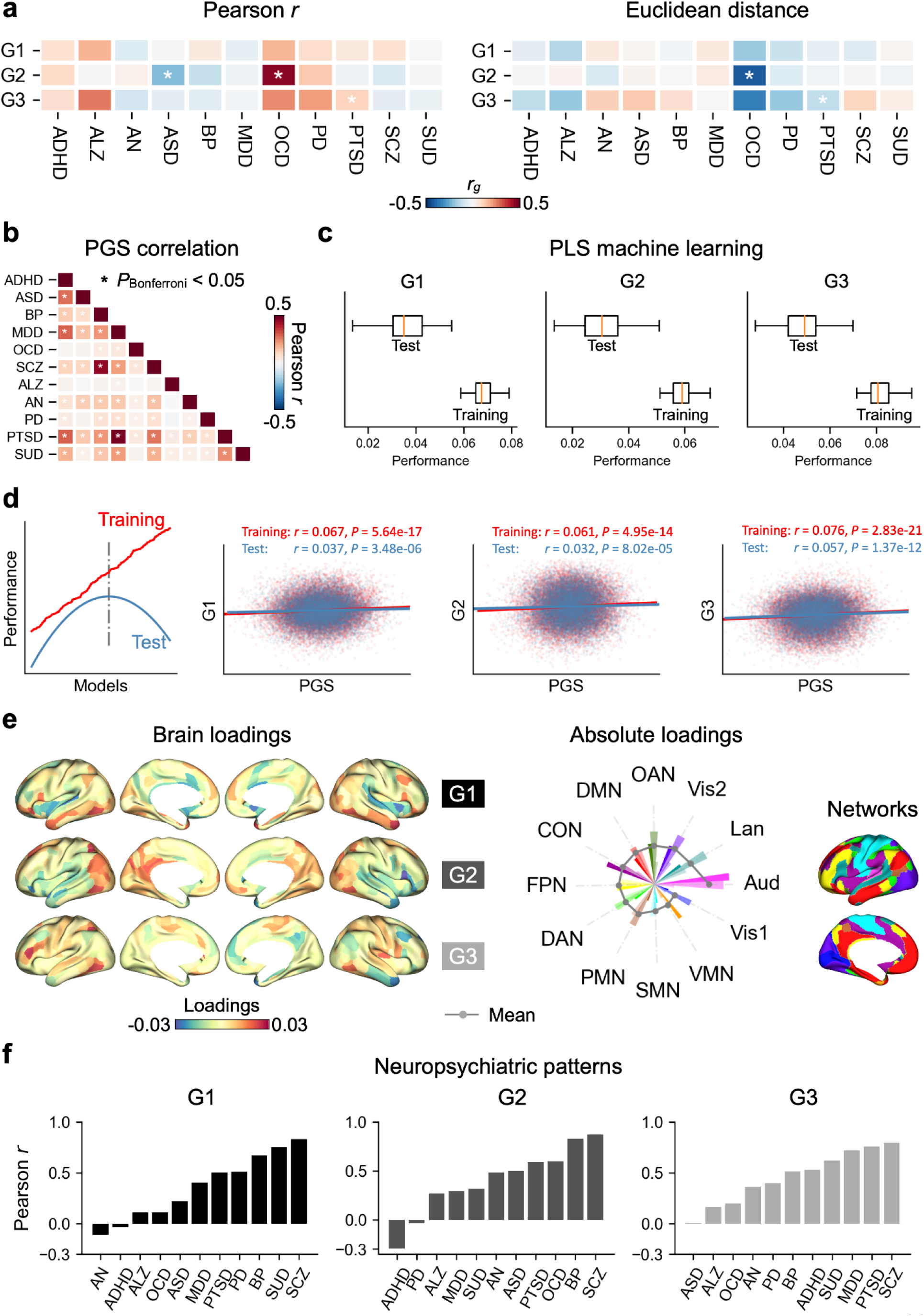
Association between regional gradients and polygenic scores for 11 neuropsychiatric conditions. **a** Shows genetic correlations between 11 neuropsychiatric conditions and Pearson *r* and Euclidean distance features. * and ** indicate *P* < 0.05 and *P*_FDR_ < 0.05. **b** Correlation between polygenic risk scores (PGSs). The 11 psychiatric conditions include attention-deficit/hyperactivity disorder (ADHD), Alzheimer’s disease (ALZ), anorexia nervosa (AN), autism spectrum disorder (ASD), bipolar disorder (BP), major depression disorder (MDD), obsessive-compulsive disorder (OCD), Parkinson’s disease (PD), post-traumatic stress disorder (PTSD), schizophrenia (SCZ), and substance use disorder (SUD). **c** Partial least squares (PLS) regression was used to associate multiple risks and regional gradients using machine learning. The sample was split into training and test subsamples 100 times. Performance used Pearson *r*. **d** The performance of the 100 machine learning iterations revealed a most generalizable model with minimum distance between training and test and maximum training (i.e., min{(training - test)/train}). **e** PLS brain loadings for each gradient. We also summarized them into 12 networks^33^ with the absolute loadings. The atlas-defined functional networks including primary visual (Vis1), secondary visual (Vis2), somatomotor (SMN), cingulo-opercular (CON), dorsal attention (DAN), language (LAN), frontoparietal (FPN), auditory network (AUD), default mode (DMN), posterior multimodal (PMN), ventral multimodal (VMN), orbito-affective (OAN). **f** Spatial correlation between brain loadings and regional *r*-map per neuropsychiatric condition and gradient in **Supplementary Fig. S13-15** and brain loading map in **d**.

## Discussion

Integrating genetic, transcriptomic, and neuroimaging data across over 30,000 individuals, we provide the first comprehensive characterization of the genetic architecture underlying cortical functional gradient organization. Through genome-wide association studies, we identified five genomic loci comprising 16 genes associated with global gradient patterns, with enrichment in metabolic pathways including lipid biosynthesis and energy metabolism. Heritability analyses across three independent cohorts spanning ages 9-70 years revealed modest but spatially similar genetic contributions, with higher heritability in association cortices than sensory regions. While direct genetic correlations between gradient organization and neuropsychiatric disorders were non-significant, multivariate analyses revealed that polygenic risk scores for schizophrenia, bipolar disorder, and PTSD significantly correlate with regional gradient patterns, particularly in auditory and language networks. Together, these findings establish functional gradients as moderately heritable features of brain organization, influenced by metabolic processes and sharing complex polygenic relationships with mental health vulnerability.

First, we found five genomic loci involved in shaping global gradient patterns, which are centered on chromosomes 1, 2, 10, and 11. Studying these loci, we found they contribute also to macrostructural features such as cortical thickness ^49–53^, surface area ^50,53^, and other features including morphological shape and sulcal depth ^50,53^. Regarding brain functional features, rs38191783 and rs4880259 have been consistently observed to be associated with amplitude of low-frequency fluctuations in many regions ^54^ and independent components of amplitude ^51^. As for non-brain features, they are mostly related to body weight and blood pressure ^55–64^. The full GWAS summaries of the gradients were strongly related to DWI-derived microstructural measures but weakly related to macrostructural measures using GWAS summaries from a previous study ^44^. This may indicate two things. Firstly, function is more closely reflective of microstructural integrity than macrostructure in the human brain; secondly, common genetic factors may push microstructural integrity (positive *r_g_* for fractional anisotropy and negative *r_g_* for mean diffusion, orientation dispersion, and ISOVF) and function gradients in similar directions. Of note, these DWI-derived measures are based on the mean score of the whole brain. Further regional patterns would benefit a detailed understanding of structure-function relationships, for example, to uncover the directionality of relationships (e.g., higher microstructural integrity may relate to both stronger between or within network integration, indirectly captured by gradient loadings). Phenotypic correlation studies show that microstructure and function correspond closely in sensory areas but diverge in association areas ^28,65,66^. This divergence possibly stems from different relationships between microstructure and short-versus long-distance functional connectivity patterns or from environmentally induced plasticity impacting structure and function differently ^28,67,68^. Mendelian randomization (MR) could help us better understand their directionality in the future. For example, previous work using this approach has shown that surface area unidirectionally drives the folding of the cortex ^44^. The identification of metabolic pathway genes is particularly noteworthy given emerging evidence that brain function is fundamentally constrained by energetic demands ^32,69,70^. Functional connectivity patterns may reflect optimization of metabolic efficiency, with higher-order association regions requiring greater energy investment for integrating diverse information streams. The genetic variants we identified may influence gradient organization by modulating cellular metabolism, neurotransmitter synthesis, or energy allocation across cortical hierarchies. However, the specific mechanisms linking genetic variation in metabolic pathways to large-scale functional architecture remain to be elucidated and represent an important direction for future mechanistic studies.

Unpacking from global to regional gradient loadings, we still found some overlapped SNPs. Though the global patterns were heritable, the regional heritability varied a lot. In particular, high heritability in frontal and parietal areas but low in somatosensory areas. We observed similar heritability patterns across UKB, HCP, and QTAB, even though the three datasets have different age ranges. This suggests that inter-individual variance in gradient loadings is spatially-consistently heritable across different age ranges. The degree of heritability was comparable for the two twin-based analyses and, as expected, lower for the SNP-based heritability. The modest heritability estimates warrant careful interpretation. This indicates that the majority of individual differences in gradient organization reflect environmental influences, developmental experiences, measurement error, or gene-environment interactions not captured by SNP-based heritability. The higher heritability in association cortices may reflect several non-mutually exclusive possibilities: (1) stronger genetic constraints on higher-order cognitive functions that emerge later in development; (2) greater environmental plasticity in sensory systems exposed to direct experience-dependent modification; or (3) technical factors such as lower signal-to-noise ratio in subcortical and sensory regions. Longitudinal studies with detailed environmental measures are needed to understand how genetic and environmental contributions interact across development ^71,72^.

Our analyses revealed important methodological challenges in imaging genetics research. Most notably, we observed no overlap between genes identified through GWAS (based on individual differences across 30,000+ individuals) and those identified through data-driven spatial correlation with AHBA gene expression (based on group-level patterns across ∼300 cortical samples from 6 donors). This discordance likely reflects fundamental differences in what these approaches measure: GWAS captures genetic variants influencing individual differences in gradient organization through causal developmental or physiological pathways, whereas AHBA spatial correlation identifies genes whose expression topography matches gradient patterns without establishing causality or relevance to inter-individual variation. The GWAS-AHBA discordance is further complicated by developmental expression differences, where G2-related genes showed opposite trajectories between GWAS-informed and AHBA-driven approaches. These findings underscore that spatial gene expression patterns, while informative about molecular organization, do not directly translate to genetic influences on individual differences. Future work integrating single-cell transcriptomics, developmental trajectories, and individual-level genetic data may help bridge these methodological divides. Additionally, rigorous preprocessing and quality control of AHBA data, including handling missing values, donor-specific effects, and hemispheric asymmetries, remains critical for robust inference ^7^.

The relationship between functional gradients and psychiatric conditions revealed a nuanced genetic architecture. Direct genetic correlations between gradient GWAS and psychiatric disorder GWAS were non-significant, indicating that the specific genetic loci influencing gradient organization do not substantially overlap with those conferring psychiatric risk. However, multivariate polygenic risk score analyses revealed significant associations, particularly for schizophrenia, bipolar disorder, and PTSD. Here SCZ is also the most heritable of disorders ^46^. This apparent contradiction suggests that gradients and psychiatric disorders share polygenic influences distributed across many small-effect variants, rather than specific shared loci. The prominence of language and auditory regions in the gradient-PGS associations is intriguing. These areas show high heritability and serve as convergence zones integrating sensory, cognitive, and emotional information ^73,74^, functions commonly disrupted across psychiatric conditions.

Language-related areas may represent vulnerability markers given their genetic links to social cognition and their integrative role spanning multiple cognitive domains. The cortical patterns we observed, distinguishing inferior frontal and middle temporal areas from sensory cortices for G1, insular areas from visual cortices for G2, and default mode from frontoparietal networks for G3, suggest that psychiatric genetic risk manifests through altered hierarchical organization rather than localized dysfunction. Importantly, gradient alterations have been observed across multiple psychiatric conditions but with inconsistent directionality ^20,21^: some studies report increased sensory-association differentiation in psychosis ^21^ while compressed in schizophrenia ^20,21^. Our PGS findings suggest these apparent inconsistencies may reflect shared vulnerability to gradient disruption rather than diagnosis-specific patterns. The transdiagnostic nature of our PGS effects aligns with growing recognition of shared genetic architectures across psychiatric conditions. Whether gradient measures could serve as intermediate phenotypes or treatment response predictors remains an important avenue for translational research.

Our findings open several critical questions for future research. First, how do metabolic pathway genes mechanistically influence functional gradient organization? Hypotheses include: modulation of synaptic energy availability affecting connection strength; influence on neurotransmitter synthesis impacting functional coupling; regulation of myelination affecting conduction velocity and temporal synchronization; or control of cellular metabolism determining neuronal firing patterns. Experimental studies manipulating metabolic pathways in model systems could test these mechanisms directly. Second, longitudinal studies are essential to understand how genetic and environmental factors shape gradient development. Do the same genetic variants influence gradients throughout life, or do different genes become relevant at different developmental stages? The consistent heritability patterns across ages 9-70 suggest stable genetic influences, but within-individual trajectories are needed to establish this definitively. Third, multi-omics integration could refine our understanding. Combining genetic data with epigenomics, proteomics, and metabolomics may reveal how genetic variation translates through regulatory networks to influence functional architecture. Single-cell transcriptomics could clarify which cell types mediate genetic influences on gradients, building on evidence that interneurons and layer-specific excitatory neurons show differential enrichment along the sensory-association axis. Finally, the clinical relevance of our findings requires further evaluation and validation. Understanding how metabolic health influences gradient organization may inform preventative strategies for cognitive decline and mental health disorders.

Several limitations warrant consideration when interpreting our findings. First, our analyses predominantly included individuals of European ancestry, limiting generalizability to other populations. Allele frequencies, linkage disequilibrium patterns, and gene-environment interactions differ across ancestries, and replication in diverse cohorts is essential. Our non-European replication sample (N = 1,444) showed consistent genomic inflation factors, suggesting similar genetic architecture, but larger samples are needed for definitive conclusions. Second, GWAS inherently focuses on common genetic variants (MAF > 1%), potentially overlooking rare variants with large effects. While common variants typically explain more population-level variance, rare variants may be critical for understanding individual cases, particularly in clinical contexts. Third, the modest heritability estimates indicate that most variance in gradient organization remains unexplained by genetic factors captured through SNP arrays. Environmental influences, including education, stress, nutrition, and early life experiences, likely contribute substantially. Gene-environment interactions and correlations, which our cross-sectional design cannot disentangle, may also be important. Future studies incorporating detailed environmental assessments and longitudinal designs are needed. Fourth, the cross-sectional design precludes causal inference. We cannot determine whether genetic variants causally influence gradients, whether gradients mediate genetic effects on cognition or psychiatric risk, or whether unmeasured confounders explain associations. Mendelian randomization approaches could help establish causality in future work. Fifth, the discordance between GWAS-informed and AHBA-driven gene sets highlights methodological limitations in integrating genetics and transcriptomics. AHBA data come from only six post-mortem donors, have incomplete cortical coverage, and lack information about individuals’ genetic backgrounds or gradient phenotypes. The spatial correlation approach assumes that gene expression topography directly relates to function, which may not hold, especially for genes with indirect or developmental effects. Sixth, all individuals’ gradients were aligned to an HCP young adult template, which may introduce age-related biases. While this ensures comparability, it assumes that the canonical gradient structure remains stable across age, potentially underestimating age-specific patterns or developmental reorganization. Finally, our focus on cortical gradients excludes subcortical structures, which show substantial individual variation and psychiatric relevance. Extending gradient analyses to whole-brain networks would provide a more complete picture of functional organization’s genetic architecture.

In conclusion, we demonstrate that cortical functional gradient organization has a complex genetic architecture involving metabolic pathway genes, shows moderate but spatially consistent heritability across the lifespan, and shares polygenic relationships with psychiatric vulnerability despite lacking specific genetic overlap. These findings establish functional gradients as biologically meaningful features of brain organization that bridge genes, metabolism, and mental health. However, the modest genetic contribution and substantial unexplained variance underscore the importance of environmental factors and gene-environment interactions in shaping functional brain architecture. Future work integrating longitudinal designs, diverse populations, multi-omics data, and mechanistic experiments will be essential to translate these findings into actionable insights for understanding and improving brain health.

## Methods

Data used in the present study are all available through their data repositories. Ethics regarding each dataset have been approved by their review committee boards. UKB data used in this study is under the application ID: 20904.

### Datasets

#### UKB

UKB is a prospective cohort of approximately 500,000 individuals from the UK, who have SNP genotype data. Of these individuals, approximately 40,000 individuals had completed the MRI scanning when the current study commenced. Participants were excluded for safety criteria such as metal implants, recent surgery, or conditions problematic for scanning such as hearing problems, breathing problems, or claustrophobia. The age range of the UKB sample was from 40 to more than 70 years. An overview of the data can be seen at the UKB website: https://www.ukbiobank.ac.uk.

#### HCP

HCP has 1,206 healthy young adults with four sessions of high-quality rs-fMRI scanning from the US ^34^. It’s a twin-based dataset with 298 monozygotic (MZ) and 188 dizygotic (DZ) twins as well as 720 singletons. We included individuals with a complete set of four fMRI scans that passed the HCP quality assessment ^34,75^. We included 470 males with a mean age of 28.7 years (range: 22–37). Data can be openly obtained from the HCP DB website: https://db.humanconnectome.org.

#### QTAB

QTAB is a longitudinal dataset including 422 subjects aged from 9 to 14 years for the baseline and 304 subjects aged from 10 to 16 years for the follow-up ^76^. Here, we used baseline data (48% female, mean age 11.3 ± 1.4 years) including 218 MZ, 194 DZ, 5 unpaired twins, and 5 without imaging data. Data can be openly obtained from OpenNeuro: https://openneuro.org/datasets/ds004146.

### Genome-wide association

Before GWAS, genetic quality control and imputation of the UKB were done by the UKB team and described in detail elsewhere ^77^. The following inclusion and exclusion criteria have been described in our previous publication ^44^. Briefly, the inclusion criteria was self-identified white ethnicity and the exclusion criteria was outliners, of the first two genetic principal components, exceeding ±5 standard deviation from the mean. These are also called predominantly European genetic ancestries. We further removed individuals whose genetic sex did not match their reported sex, or had excessive genetic heterozygosity, as provided by the UKB team. For the GWAS, we used all genotyped and imputed SNPs in the UKB with a common variant with minor allele frequency (MAF) > 1%, which were in Hardy–Weinberg equilibrium (HWE; *P* < 1 × 10^−6^) and, for imputed SNPs, had an imputation *r*^2^ > 0.4. After genetic quality control, we retained 31,797 participants (not the final sample) and approximately 9.1 million SNPs. We conducted three GWAS for the global features (described below in the functional gradients section) using fastGWA-GLMM in the GCTA toolbox (version 1.94.1) ^31^. A genetic relationship matrix (GRM) was used in fastGWA as a random variable. For all GWAS, we included age, age2, sex, age × sex, age2 × sex, imaging center, first 40 genetic principal components, and mean framewise displacement. Framewise displacement and Euler number are common covariates in fMRI studies ^23,78^ and in order to make comparisons with a previous structural GWAS study ^44^ that included them we also included them as covariates. We identify associations using the common genetic variant significance level *P* < 5 × 10^−8^.

After obtaining GWAS results, we used the Functional Mapping and Annotation (FUMA) ^36^ toolbox to map the genes. Lead SNPs were first identified, and genomic regions around each lead SNP were defined using a window size of 10 kb. SNPs were then mapped to the nearest genes, and regions were extended based on linkage disequilibrium (LD) patterns to include SNPs in LD with the lead SNPs. We performed gene-based association analysis using MAGMA implemented in the FUMA platform. MAGMA aggregates SNP-level association statistics into gene-level statistics using a multiple linear principal components regression model, while accounting for LD between SNPs. GWAS summary statistics were used as input, and SNPs were mapped to protein-coding genes based on genomic location using default MAGMA annotation, assigning SNPs to genes within gene boundaries (no additional window). LD structure was estimated using the European reference panel from the 1000 Genomes Project (Phase 3), consistent with the ancestry of our GWAS sample. For each gene, MAGMA computes a gene-level p-value based on the joint effect of all SNPs mapped to that gene. Genome-wide significance was defined using Bonferroni correction for the number of tested genes (P < 0.05 / 19,673). The resulting gene-level statistics were used to identify significantly associated genes and were compared with genes obtained from positional mapping in FUMA to assess consistency and complementarity across methods.

### Imaging preprocessing

Details on data processing and acquisition can be found in the UKB Brain imaging documentation (https://biobank.ctsu.ox.ac.uk/crystal/crystal/docs/brain_mri.pdf). Briefly, individual rs-fMRI data were motion corrected, intensity normalized, high-pass temporally filtered, and further denoised using the ICA-FIX pipeline, all implemented in FSL. Multimodal parcellation (MMP) ^30^ was warped to subject space based on the high-resolution T1-weighted anatomical images. Individual warping parameters were applied to map the parcellation to the functional space following T1-rsfMRI alignment. Finally, FC was calculated by Fisher-z transforming the correlation matrix of MMP parcellated time series. Genetic and imaging quality control resulted in 30,716 subjects (the final sample, age: 45-82 years, mean: 64 ± 7 years, 53.1% females) from UKB used for GWAS in this study. For the replication sample, we used a new independently released sample of UKB (*N* = 14,471, age: 53-87 years, mean: 70 ± 7 years, 53.9% females).

All HCP rs-fMRI data underwent HCP’s minimal preprocessing ^75^ and were coregistered using a multimodal surface matching algorithm (MSMAll) (Robinson et al., 2014) to the HCP template 32k_LR surface space. The template consists of 32,492 total vertices per hemisphere (59,412 excluding the medial wall). We calculated the correlation matrix of MMP parcellated time series for two sessions (REST1_LR and REST1_RL). Then, we averaged the correlation matrix and Fisher-z transformed it to obtain the FC matrix. Finally, we analyzed 1,018 subjects.

For the QTAB, we followed the published pipeline ^79^. Briefly, individual rs-fMRI data were motion and distortion corrected, high-pass temporally filtered, and then nuisance signals removed. The native surface was reconstructed from the T1w image using fastsurfer. Processed time series data were then registered to the native cortical surface and further registered to fsLR_32k space with a 10mm Gaussian kernel smooth. We calculated the FC matrix of MMP parcellated time series for 422 subjects.

### Functional gradients

Next, we employed the nonlinear dimensionality reduction technique ^3^ to generate the gradients of the FC matrix. We first made the FC matrix sparse with the top 10% of the connectivity remaining, then applied diffusion embedding on the affinity matrix, and finally extracted the first 10 eigenvectors with 360 eigenvalues for each eigenvector. All the steps were accomplished using the Python package BrainSpace ^10^. Along these low-dimensional axes or gradients, cortical regions with similar functional connectivity profiles are situated together. The name of this approach, which belongs to the family of graph Laplacians, is derived from the equivalence of the Euclidean distance between points in the diffusion-embedded mapping ^3,10,80^. It is controlled by a single parameter α, which controls the influence of the density of sampling points on the manifold (α = 0, maximal influence; α = 1, no influence). On the basis of the previous work ^3^, we followed recommendations and set α = 0.5, a choice that retains the global relations between data points in the embedded space and has been suggested to be relatively robust to noise in the covariance matrix.

Group-level gradients were constructed using the HCP averaged FC matrix and individual gradients were generated using Procrustes alignment from raw gradients to HCP group-level gradients (template), which makes individuals comparable. The global features are the similarity value (Pearson *r and* Euclidean distance) between the individual gradients after alignment and template gradients. The regional features are the regional eigenvalues after alignment for each individual. Here, we conducted the analyses for the first three gradients including: sensory-association, visual-somatomotor, and modulation-representation.

At the same time, we used the UKB group-level gradients as the template to generate the individual gradients. It showed a high correlation with HCP as the template (*r*_G1_ = 0.95, *r*_G2_ = 0.97, and *r*_G3_ = 0.88). Their genetic correlations are also high (*r*_g_G1_ = 0.62, *r*_g_G2_ = 0.90, and *r*_g_G3_ = 0.70, **Supplementary Fig. S7**).

### Transcriptomics

We used BrainSpan and AHBA to investigate how the gradient gene expression in the brain develops over age and whether regional expression is correlated with gradient maps. BrainSpan contains transcriptomics of the 29 developing brain samples from 8 post-conceptual weeks (PCW) to 40 years old, data offered from FUMA ^36,81^ (https://fuma.ctglab.nl/). AHBA contains microarray expression data from approximately 20,000 genes ^38^. A total of 3,702 brain tissue samples were collected from six neurotypical adults, including three European males, two African American males, and one European female. ENIGMA-toolbox ^41^ integrates the gene expression data from the abagen ^40^ and helps project the averaged gene expression data from the Desikan–Killiany atlas to MMP, where 15,634 genes on 312 parcels are available to obtain the transcriptomic data. We imputed the missing values using the variogram algorithm based on the central coordinates of the parcels and used imputed data to do the transcriptomic analyses (full 360 parcels). The toolbox we used here is PyKirge (https://pykrige.readthedocs.io).

After obtaining the genes, we performed KEGG enrichment analysis using ShinyGO 0.81 ^45^. Edges represent similarity between pathways, based on the proportion of shared genes between two pathways. The equation = ∣A∪B∣/∣A∩B∣, where A and B are the sets of genes in two different pathways. In ShinyGO, a similarity cutoff (> 0.3) is used to determine whether to draw an edge between two pathways.

### Heritability analyses

For the SNP-based heritability, we used GCTA–GREML (v1.94.1) ^31,82^ to calculate how much the phenotypic variance is explained by all the SNPs. We included age, age2, sex, age × sex, age2 × sex, imaging center, first 40 genetic principal components, mean framewise displacement, maximum framewise displacement, and Euler Index as covariates. For the twin-based heritability, we used Sequential Oligogenic Linkage Analysis Routines (SOLAR, v9.0) ^83^. Heritability (i.e. narrow-sense heritability *h*^2^) represents the proportion of the phenotypic variance (σ^2^_p_) accounted for by the total additive genetic variance (σ^2^_g_), that is *h*^2^ = σ^2^_g_ / σ^2^_p_. Phenotypes exhibiting stronger covariances between genetically more similar individuals (MZ twins) than between genetically less similar individuals (other pairs) have higher heritability. We added covariates to our twin-based heritability models including age, sex, age2, and age×sex. The spatial correlations between datasets across gradients were corrected using spatial autocorrelation ^84^, which generates surrogate maps using the variogram function on the geometric distance matrix. We generated 1000 surrogate maps as null models and tested how many null models the practical model exceeded to obtain the *P*_variogram_ value.

### PGS for neuropsychiatry conditions

We calculated the PGS for 11 psychiatric conditions including: ADHD ^85^, ALZ ^86^, AN ^87^, ASD ^88^, BP ^89^, MDD ^90^, OCD ^91^, PD, PTSD ^49^, SCZ ^92^, and SUD ^93^. Summary statistics were used in independent cohorts from the UKB, filtered for common variants also present in the UKB cohort in our GWAS. PGS weights were derived for each chromosome separately, using PRS by continuous shrinkage (PRS-cs) ^94^, based on the LD reference panel for predominantly European ancestry from the 1000 Genomes Project. Each individual was scored for each chromosome and the total PGS across 23 chromosomes was standardized to z-scores (zero mean, unit variance). We used published GWAS summary statistics for 13 imaging-derived phenotypes of global brain macro- and microstructure ^44^. For these phenotypes, we estimated their genetic correlation with the functional gradients from the present study using LD score regression (LDSC) ^42,43^. The same method was also applied to estimate the genetic correlation amongst the three functional gradients, and their heritability.

### PLS machine learning

We initially conducted PLS analyses with a machine learning pipeline to uncover the overall multivariate association between regional gradients (360 x 30,716) and PGSs (11 x 30,716 x 3 gradients). We regressed out covariates for regional gradients including age (age at scanning, age^2^ at scanning, and time gap between age at scanning and blood test), sex (genetic sex, sex*age, and sex*age^2^), scanning site, and head motion (Euler angles for left plus right hemispheres, mean framewise displacement, and maximum framewise displacement), and first forty genetic components. Regarding the machine learning pipeline, we split the subjects into training and test samples 100 times and performed PLS in the training sample and transformed the test sample to look at the brain-PGS first dimension’s correlation. We also tested the GRM diagonal differences between the two splits for every iteration, which showed no significant difference. Then, we selected the best model as the output, which maximally utilizes performance in the test sample and reduces the overfitting outcome, i.e., minimum distance between training and test and maximum training (min{(training - test)/train}. To capture the three gradients at the same time, we used the minimal averaged performance across three gradients as the final model. We focused on the first component of neuropsychiatric conditions and brain features for three gradients separately. After obtaining the latent brain loadings for each gradient, we calculated the Pearson *r* between brain loadings and each neuropsychiatric PGS effect size map. Finally, we were able to determine how similar each neuropsychiatric PGS was to the latent brain loadings.

### Multiple comparisons correction

In **Fig. 1**, we used a classical threshold (5 × 10^−8^) for the global feature of GWAS summaries. In **Fig. 2**, we used the threshold (5 × 10^−8^)/360 for the regional GWAS. In **Fig. 3**, considering the spatial autocorrelation between two brain maps, we simulated 1000 surrogate brain maps using BrainSMASH ^84^ using the variogram algorithm based on geodesic distance of Glasser parcellation. The P_variogram_ was calculated to control spatial autocorrection by how many correlations the practical map [test and target] is greater than the 1000 surrogate maps [test and surrogate]. In **Fig.4**, multiple linear regression with P < 0.05 for each model was used. **In Fig. 5**, we used FDR correction for the LDSC genetic correlation between our GWAS summaries and published brain structural GWAS summaries. **In Fig. 6**, no statistical tests were used. **In Fig. 7**, genetic correlations between GWAS summaries were controlled by FDR and correlations between PGS scores were controlled by Bonferroni correction. In addition, we used the uncorrected P-values for the PLS brain-PGS association.

## Data availability

The raw data we used in the current study are available through their respective websites, which has been mentioned in the Datasets section. Source data is also provided.

## Code availability

All scripts including genetic analyses, brain features computation, and statistical models can be found at a GitHub repository: https://github.com/wanb-psych/GWAS_FuncGradients.

## Acknowledgments

BW is supported by the International Max Planck Research School on Neuroscience of Communication: Function, Structure, and Plasticity (IMPRS NeuroCom), the Federation of European Neuroscience Societies (FENS), and the IBRO Pan-Europe Regional Committee (IBRO-PERC) Exchange Fellowships Programme. SLV is funded by the Otto Hahn Award and Lise Meitner Excellence Program at the Max Planck Society and Helmholtz International BigBrain Analytics and Learning Laboratory (HIBALL), supported by the Helmholtz Association’s Initiative and Networking Fund and the Healthy Brains, Healthy Lives initiative at McGill University.

## Authors’ contribution

Conceptualization: B.W., S.L.V. Methodology: B.W., W.V, R.A.I.B, S.L.V. Formal analysis: B.W., Y.H. Writing - Original Draft: B.W., S.L.V. Writing - Review & Editing: B.W., H.Y., W.V., A.J., S.B.E, R.A.I.B., S.L.V. Visualization: B.W. Data curation: B.W., R.A.I.B, S.L.V. Project administration: B.W. Funding acquisition: B.W., S.L.V. Supervision: S.L.V.

